# Globus pallidus externus (GPe) alpha band activity decreases after deep brain stimulation in clinically responsive obsessive-compulsive disorder patients

**DOI:** 10.64898/2026.04.10.26350428

**Authors:** Zarghona Imtiaz, Brian H. Kopell, Sonia Olson, Ignacio Saez, Ha Neul Song, Helen S. Mayberg, Ki Sueng Choi, Allison C. Waters, Martijn Figee, Andrew H. Smith

## Abstract

**Background:** Deep brain stimulation (DBS) of the anterior limb of the internal capsule (ALIC) is an effective treatment for severe obsessive-compulsive disorder (OCD). Identifying brain readouts of positive response may guide further DBS optimization.

**Methods:** We measured local field potential (LFP) changes from bilateral DBS leads in 10 OCD patients implanted at a uniform tractographic network target derived from prior DBS responders. We consistently stimulated dorsal lead contacts in the ALIC white matter, while recording LFP from the ventral lead contacts in grey matter of the anterior globus pallidus externus (GPe), a key node in the basal ganglia non-motor indirect pathway.

**Results:** After six months of DBS, OCD symptoms decreased on average by 40% across subjects, along with a significant decrease in alpha activity across both hemispheres. Only one patient did not have an improvement of symptoms, and this was also the only patient to never exhibit an alpha decrease in either hemisphere.

**Conclusions:** Our findings suggest that therapeutic ALIC DBS coincides with a stable decrease in limbic-cognitive GPe alpha power, which should be further investigated as a potential biomarker of sustained response.

## Introduction

Obsessive-compulsive disorder (OCD) is characterized by recurring intrusive thoughts as well as repetitive behaviors, causing substantial distress that can interfere with daily life. Deep brain stimulation (DBS), chronically implanted brain electrodes delivering high frequency electrical pulses to a target region, is an effective and accepted treatment for refractory OCD (U.S. Food & Drug Administration, 2009). However, DBS for OCD shows variable outcomes likely reflecting variable neural activity in targeted brain regions. One proposed brain target for OCD has been the corticostriatal direct (‘go’) pathway which, when overactive, may drive repetitive compulsive behaviors (Ahmari & Rauch, 2022). However, DBS in the subthalamic nucleus (STN), an important node of the opposing indirect (‘no go’) corticostriatal pathway, has excellent anti-OCD effects (Mallet et al., 2008). Further supporting the indirect pathway as a target for OCD, effective DBS in the anterior limb of the internal capsule (ALIC) involves white matter fibers connected to the anterior (limbic) portions of STN (Li et al., 2020; Smith et al., 2021), as well as STN-GPe indirect pathway connections (Hollunder et al., 2024). Using prospective tractographic analyses of ALIC DBS for OCD, we also recently confirmed the therapeutic relevance of these indirect pathway fibers (Segura-Amil et al., 2025). Finally, inhibition of GPe indirect pathway projections reduced compulsivity in an animal model of OCD (Piantadosi et al., 2024). In sum, compelling cross-species evidence now points to the indirect pathway, modulation of which may free patients up to think and act in a less restrained and less perseverative manner (Smith, 2023).

To date, however, only very limited recordings have been obtained from indirect pathway nodes in the setting of OCD DBS. Increased theta oscillations in the anteromedial STN was demonstrated in a single patient with STN DBS for OCD relative to patients with STN DBS for Parkinson’s Disease (Rappel et al., 2018), and STN beta oscillations were shown in patients with OCD or Parkinson during a stopping task (Bastin et al., 2014). Taking advantage of newly available commercial DBS devices that are capable of stimulating and sensing, we examined longitudinal changes in GPe neural activity following ALIC DBS in patients with OCD to identify a potential biomarker of DBS effectiveness.

We examined GPe LFP activity in each frequency band at baseline (before activating DBS) and at the conclusion of 6 months of active treatment (Figure 1). Effective DBS treatment coincided with overall decrease in GPe alpha power (7-12 Hz). However, this electrophysiological pattern was not observed in our one case of complete clinical non-response. Examination of alpha oscillations across available intermediate time-points confirmed that this clinical non-responder was the only individual whose alpha power never decreased, indicating that changes in GPe alpha power may constitute a novel readout of OCD DBS response.

**Figure 1:**
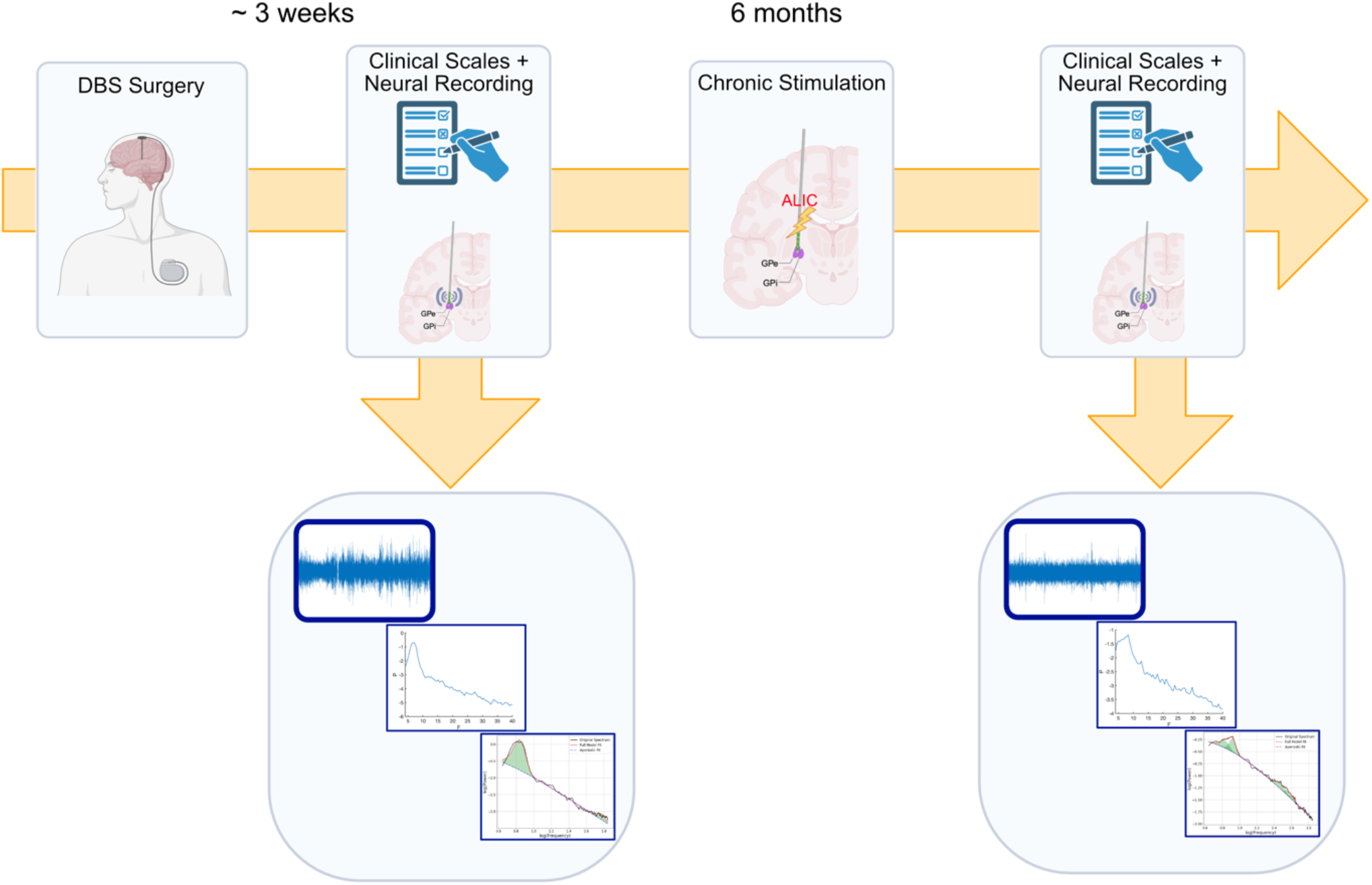
Illustration of Data Collection Design. Approximately 3 weeks after the complete deep brain stimulation (DBS) system has been implanted, a Yale-Brown Obsessive Compulsive Scale (Y-BOCS) is administered. On the same day, a 5-minute resting state eyes-open recording is performed in the globus pallidus externus (GPe). Raw time domain data from this neural recording is transformed into a power spectrum and then run through the ‘fitting oscillations & one over f’ FOOOF pipeline to derive amplitude values for periodic activity in each of the canonical frequency bands. After 6-months of active stimulation, all procedures were repeated in all patients.

## Methods

All research was performed in accordance with approval from the Icahn School of Medicine at Mount Sinai Institutional Review Board (Study #21-00125), and informed consent was obtained from all patients. Demographics and clinical characteristics are shown in Table 1. All patients underwent DBS surgery as per our standard clinical protocol, introduced above. In each patient, the location of dorsal lead contacts was selected based on individual white matter tractography, in order to stimulate the ALIC at the junction of specific white matter pathways implicated in our previous OCD DBS responders. These pathways consisted of projections between STN, brainstem, thalamus, and orbitofrontal cortex (OFC) and ventrolateral prefrontal cortex (vlPFC) (Chamberlain et al., 2008; Segura-Amil et al., 2025; van den Heuvel et al., 2005). Consistent surgical placement of the dorsal lead contacts for therapeutic stimulation of this white matter responder map was facilitated by consistent anatomical anchoring of the ventral lead contacts in the anterior (limbic-cognitive) portion of the GPe grey matter. This two-pronged approach allows delivery of on-target therapeutic stimulation to critical white matter bundles (dorsal contacts), while simultaneously enabling LFP recording in the GPe (ventral contacts), a critical grey matter node (Figure 2). Intraoperatively, prior to placing DBS electrodes, the neurosurgeon (B.H.K.) temporarily inserted a microelectrode (Alpha-Omega, Nazareth, Israel) to confirm the position of the ventral ‘anchor spot’ within the GPe grey matter. After this confirmation, the permanent SenSight DBS leads (Medtronic, Minnesota, USA) were implanted. Leads in each patient were subsequently connected to a single Medtronic Percept pulse generator (IPG) in the right chest, a device designed to both provide stimulation and record LFPs.

**Table 1:**
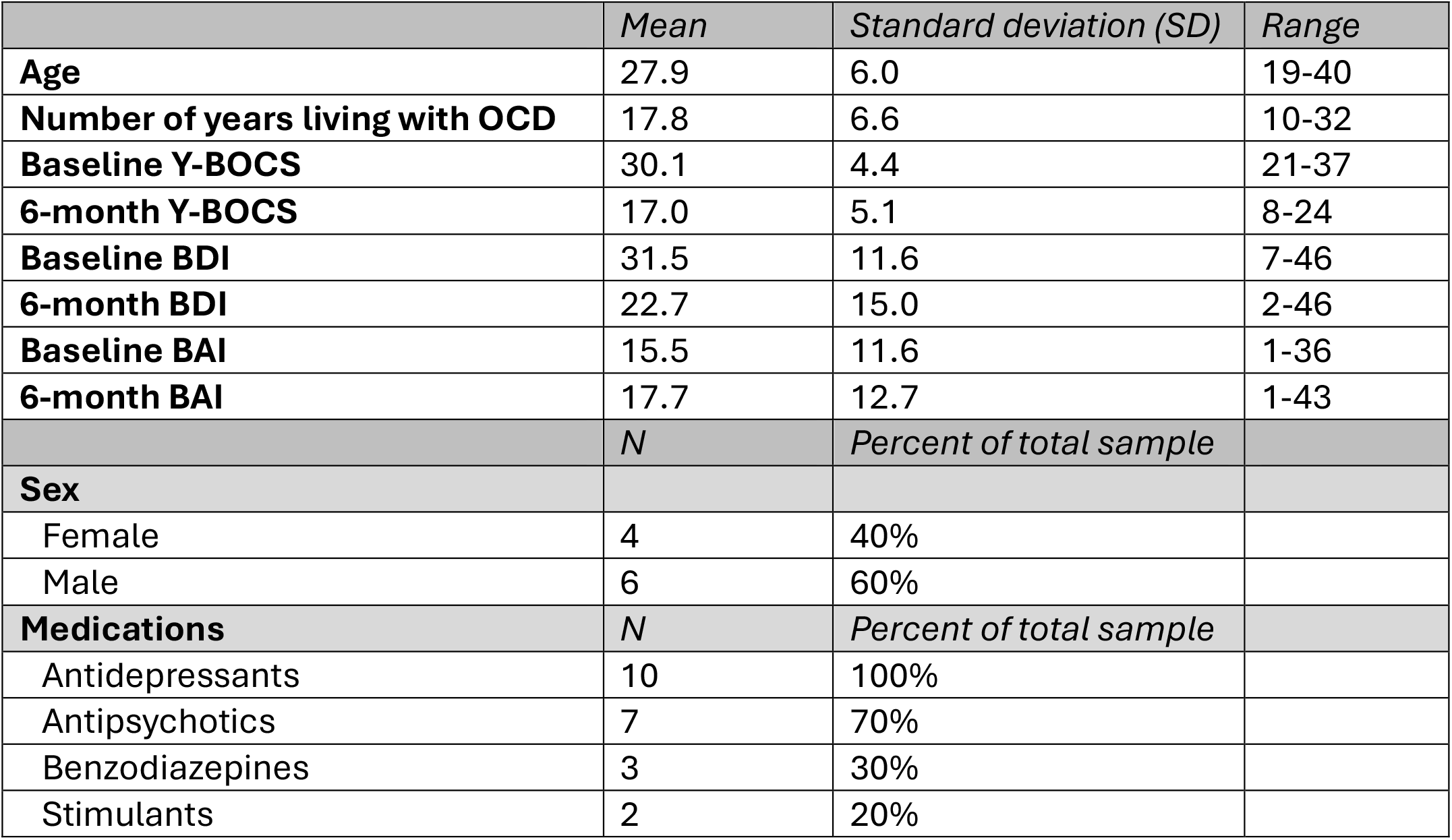
Cohort Demographics and Clinical Characteristics. Obsessive-Compulsive Disorder (OCD); Yale-Brown Obsessive Compulsive Scale (Y-BOCS); Beck Depression Inventory (BDI); Beck Anxiety Inventory (BAI).

|  | *Mean* | *Standard deviation (SD)* | *Range* |
| --- | --- | --- | --- |
| **Age** | 27.9 | 6.0 | 19-40 |
| **Number of years living with OCD** | 17.8 | 6.6 | 10-32 |
| **Baseline Y-BOCS** | 30.1 | 4.4 | 21-37 |
| **6-month Y-BOCS** | 17.0 | 5.1 | 8-24 |
| **Baseline BDI** | 31.5 | 11.6 | 7-46 |
| **6-month BDI** | 22.7 | 15.0 | 2-46 |
| **Baseline BAI** | 15.5 | 11.6 | 1-36 |
| **6-month BAI** | 17.7 | 12.7 | 1-43 |
|  | *N* | *Percent of total sample* |  |
| **Sex** |  |  |  |
| Female | 4 | 40% |  |
| Male | 6 | 60% |  |
| **Medications** | *N* | *Percent of total sample* |  |
| Antidepressants | 10 | 100% |  |
| Antipsychotics | 7 | 70% |  |
| Benzodiazepines | 3 | 30% |  |
| Stimulants | 2 | 20% |  |

**Figure 2.**
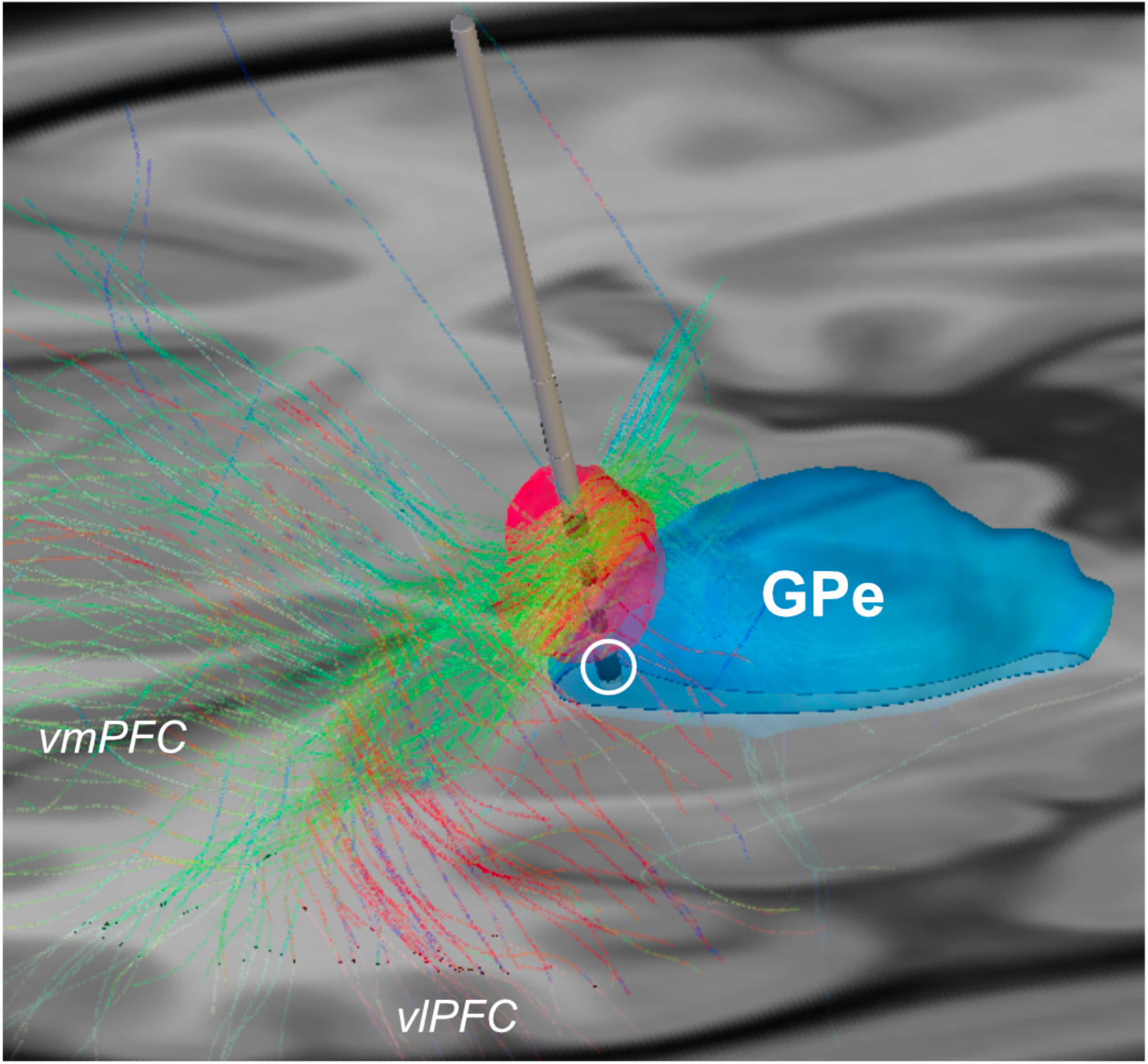
Deep Brain Stimulation Lead Placement. Anterolateral view of a sample deep brain stimulation (DBS) lead (grey, with each of the four contacts represented by dark coloring). The ventral most contact (white circle) is anchored in the anterior globus pallidus externus (GPe, blue) grey matter, where local field potential (LFP) recordings were performed. Dorsal electrode contacts are located within the anterior limb of the internal capsule (ALIC) white matter for therapeutic stimulation of axonal fibers between frontal cortex and midbrain. The volume of tissue activated (VTA) is shown in bright red around the stimulation location. Fibers are color coded based on their projection direction: green coloring represents anterior/posterior direction, blue coloring superior/inferior, and red coloring medial/lateral. Fibers terminate in ventromedial prefrontal cortex (vmPFC) and ventrolateral prefrontal cortex (vlPFC).

At approximately 1-month post-surgery, and before initiation of therapeutic stimulation, we administered a pre-treatment Yale-Brown Obsessive Compulsive Disorder Scale (Y-BOCS) and performed a baseline GPe recording. Patients were instructed to sit comfortably and quietly, with eyes open, for five minutes. Recordings of broad spectrum LFP data were obtained using the Brain Sense Streaming function of the Medtronic Percept. LFPs were captured from the ventral-most electrode contact in the anterior GPe. At the end of this appointment, the device was activated at the predefined optimal contact. In all patients, post-operative lead position had been confirmed via by a post-operative CT scan. Over the subsequent six months, minimal DBS setting adjustments were required, and when made were based on clinical judgement of the treating psychiatrist. At six months, all patients underwent a repeat Y-BOCS and second GPe recording session, performed in the same room according to the same procedures. Patients were also encouraged to present for in-person bimonthly visits. Whenever possible, symptom and electrophysiological assessments were also collected at these ‘intermediate’ timepoints between 0 and 6 months.

To examine the pre- and post-treatment LFP recordings, we used a short-time Fourier transform to convert the time domain signal into a time-frequency domain signal and plotted the power spectrums (Neumann et al., 2021). We then ran the data through the ‘fitting oscillations & one over f’ algorithmic pipeline (FOOOF) to remove the ‘aperiodic’ (i.e., non-specific) component, thereby isolating activity in specific frequency bands (Donoghue et al., 2020). FOOOF was used to best fit the data within each patient and each hemisphere, however the choice of model fit parameters was kept consistent across timepoints to facilitate cross-timepoint comparisons. The frequency range of 4-70 Hz and minimum peak height of 0.05 were kept consistent across all patients and hemispheres. Using the post-FOOOF signal, we computed the area under the curve (AUC) in the different canonical frequency bands, defined as theta (4-7 Hz), alpha (7-12 Hz), beta (12-30 Hz), and low gamma (30-70 Hz) and compared the AUC at the device activation appointment and the 6-month follow-up appointment. We pooled data from left and right hemispheres and also examined them separately, comparing timepoints using paired t-tests followed by stringent Bonferroni correction for multiple comparisons. Based on our findings in the alpha range when comparing pre- to post-treatment timepoints, we also examined alpha power in the available ‘intermediate’ timepoints between 0 and 6 months. Intermediate data were examined using a linear mixed effect model accounting for hemisphere and repeat measures.

## Results

Ten patients received ALIC DBS surgery for OCD as described above and underwent GPe recording sessions that generated broad spectrum time domain LFP data at both the baseline and six-month timepoints. Our clinical implantation strategy involves stimulating the ALIC at the confluence of specific white matter pathways involved in our previous OCD DBS responders (Segura-Amil et al., 2025). To facilitate this implantation trajectory, the ventral most portion of the DBS lead is anchored in GPe grey matter, which in turn allows us to record high-quality LFPs.

The average starting Y-BOCS for this cohort of patients was 30.1±4.4 (mean +/-standard deviation (SD)), and the average post-treatment Y-BOCS at 6 months was 17.0±5.1 (mean +/-SD). The average Y-BOCS decrease was 42.9% (paired t-test; t(9) = 7.1, 95% CI = [8.9 17.3], p= 5.8x10^-5^) (Figure 3A). In contrast, neither the Beck Depression Inventory (BDI) (paired t-test; t(8) = 1.9, 95% CI = [-1.6 16.1], p=0.09) nor the Beck Anxiety Inventory (BAI) scores (paired t-test; t(8) = -0.8, 95% CI = [-10.4 4.8], p= 0.4) showed a substantial change after 6 months of treatments, suggesting the clinical effect was specific to OCD symptoms and not depression or anxiety.

**Figure 3.**
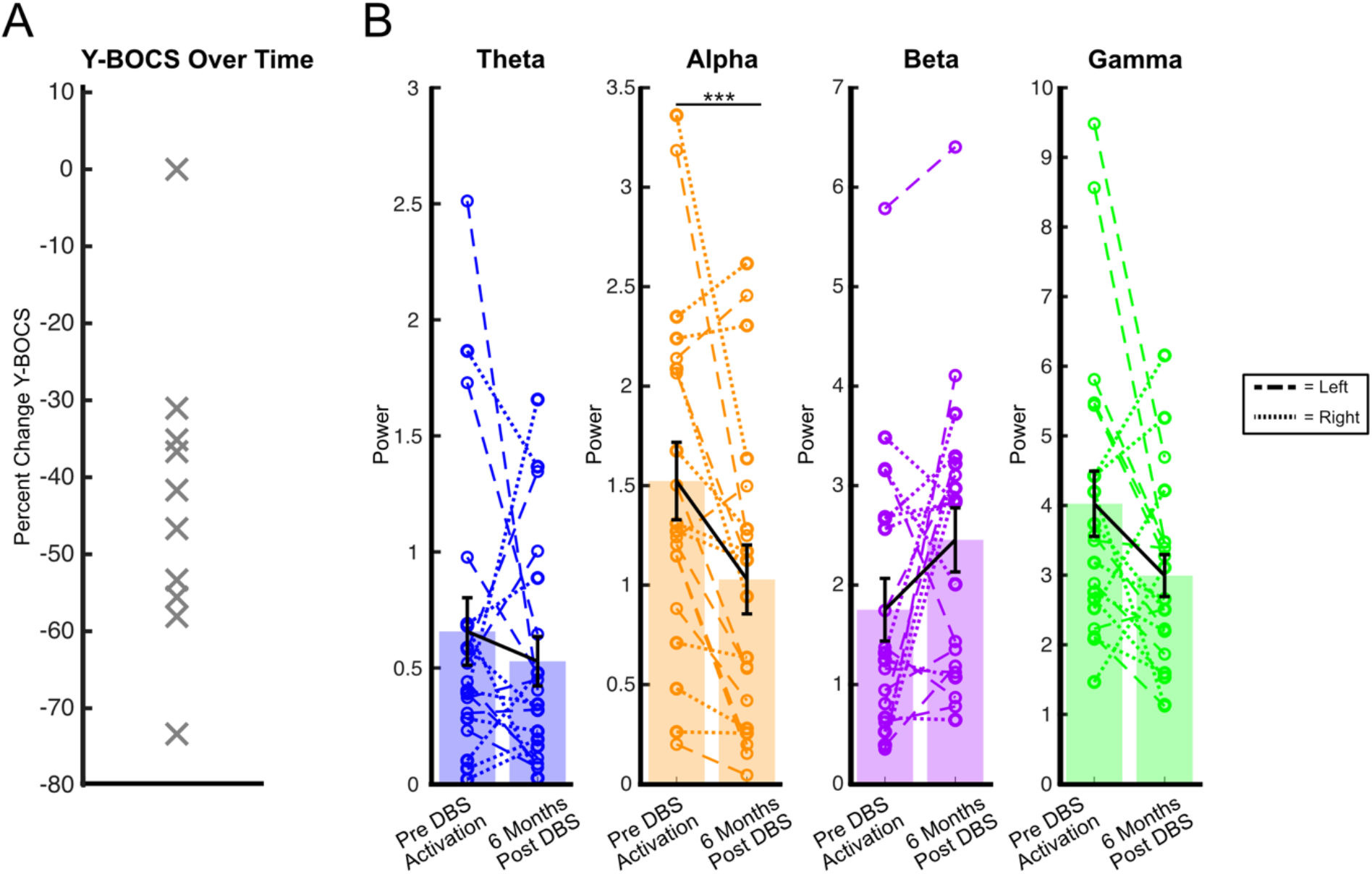
Clinical and Electrophysiological Changes in the Patient Cohort. **A**) Percent change in Yale-Brown Obsessive Compulsive Scale (Y-BOCS) after 6 months in 10 subjects. **B**) Power in each canonical frequency band prior to device turn on, and 6 months after device turn on. Data are shown from the left globus pallidus externus (GPe) (dashed lines) and right GPe (dotted lines). Mean values are indicated by black line, and error bars reflect standard error of the mean. After correction for multiple testing, only the alpha band decrease was significant (p=2.8x10^-3^, paired t-test; all others corrected p>0.05, paired t-tests). ***p<0.005

Comparing the AUC of LFP power in each frequency band between baseline and 6-month, we found a significant decrease in alpha activity (paired t-test; t(19) = 3.4, 95% CI = [0.19 0.80], p= 2.8x10^-3^) (Figure 3B). When examining hemispheres separately, alpha decreases were found in both hemispheres but were only significant in the left hemisphere (right hemisphere alpha: paired t-test; t(9) = 1.8, 95% CI = [-0.09 0.78], p = 0.1; left hemisphere alpha: paired t-test; t(9) = 3.0, 95% CI = [0.16 1.12], p = 0.01). Only one patient did not exhibit a Y-BOCS decrease after 6 months of DBS, and in this patient we found an increase in bilateral alpha at 6 months rather than a decrease (Figure 4). Removing this single patient with no clinical improvement resulted in a larger alpha decrease between baseline and 6 months (t(17) = 4.0, 95% CI = [0.27 0.89], p = 9.3x10^-4^).

**Figure 4.**
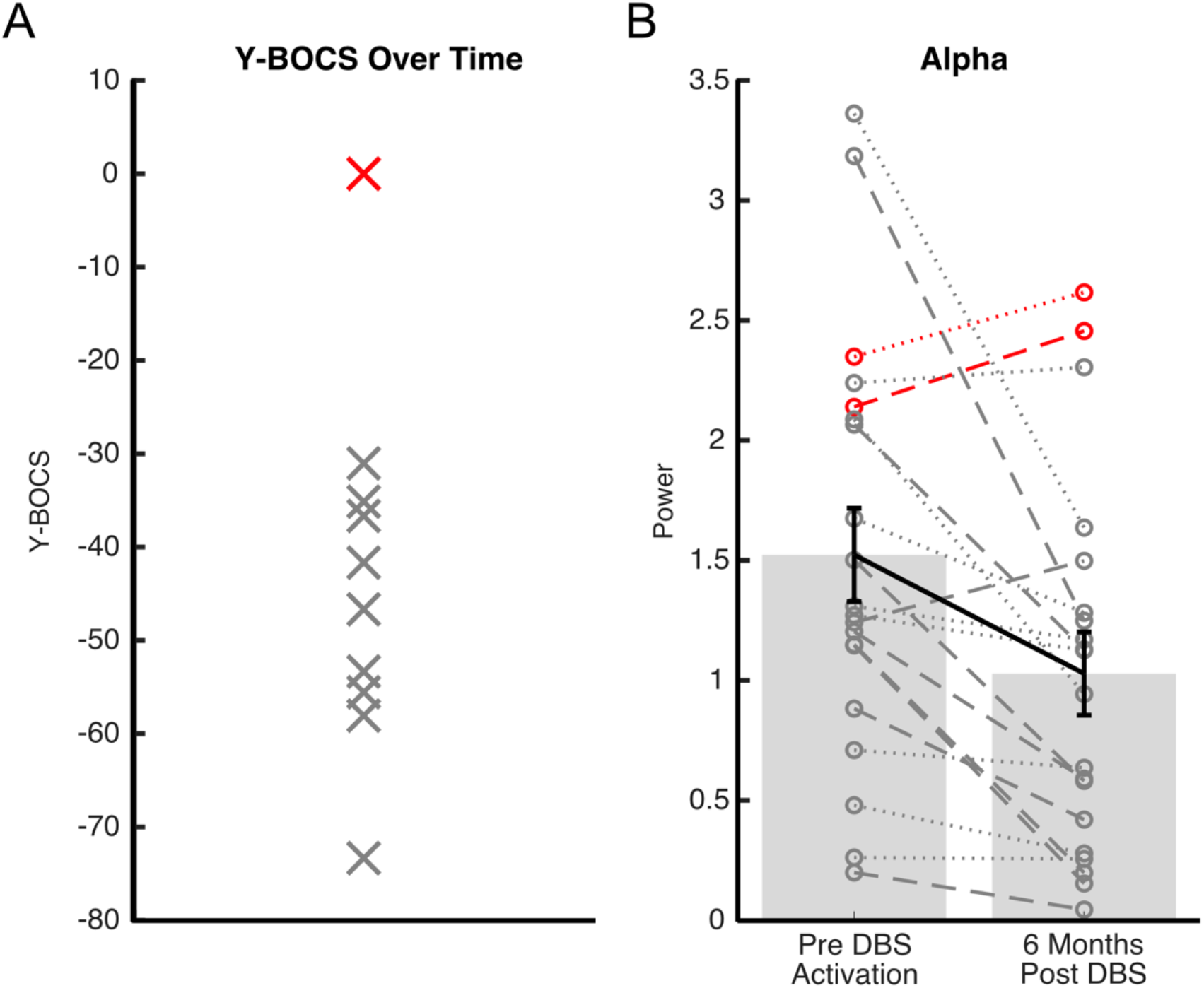
Electrophysiological Data in the Case of Deep Brain Stimulation Complete Non-Response. **A**) Percent change in the Yale-Brown Obsessive Compulsive Scale (Y-BOCS) after 6 months of active simulation as shown in Figure 3, except highlighted in red is the individual who had no change in their Y-BOCS score after treatment. **B)** Alpha band power in globus pallidus externus (GPe), with the same individual highlighted in red, showing they were the only subject to not exhibit alpha decrease in either hemisphere, instead uniquely showing bilateral alpha increase.

To further investigate longitudinal alpha dynamics, we examined available data from intermediate timepoints between 0 and 6 months (85% of bimonthly timepoints between 0 and 6 months had clinical and neural data) (Figure 5). Analyzing alpha power across all available timepoints, we found a significant effect of time on change in alpha power (linear mixed effect model; t(52) = - 3.1, p = 3x10^-3^). When adding Y-BOCS to the model there was a significant relationship between the time course of alpha power change and the time course of Y-BOCS change (linear mixed effect model; t(51) = 2.1, p = 0.04). The single patient whose Y-BOCS did not decrease after 6 months was noted to never have exhibited a Y-BOCS decrease during the six months, and was also noted to be the only patient who never exhibited any alpha decrease in either hemisphere during the six months (Figure 5A: alpha power; Figure 5B: Y-BOCS).

**Figure 5.**
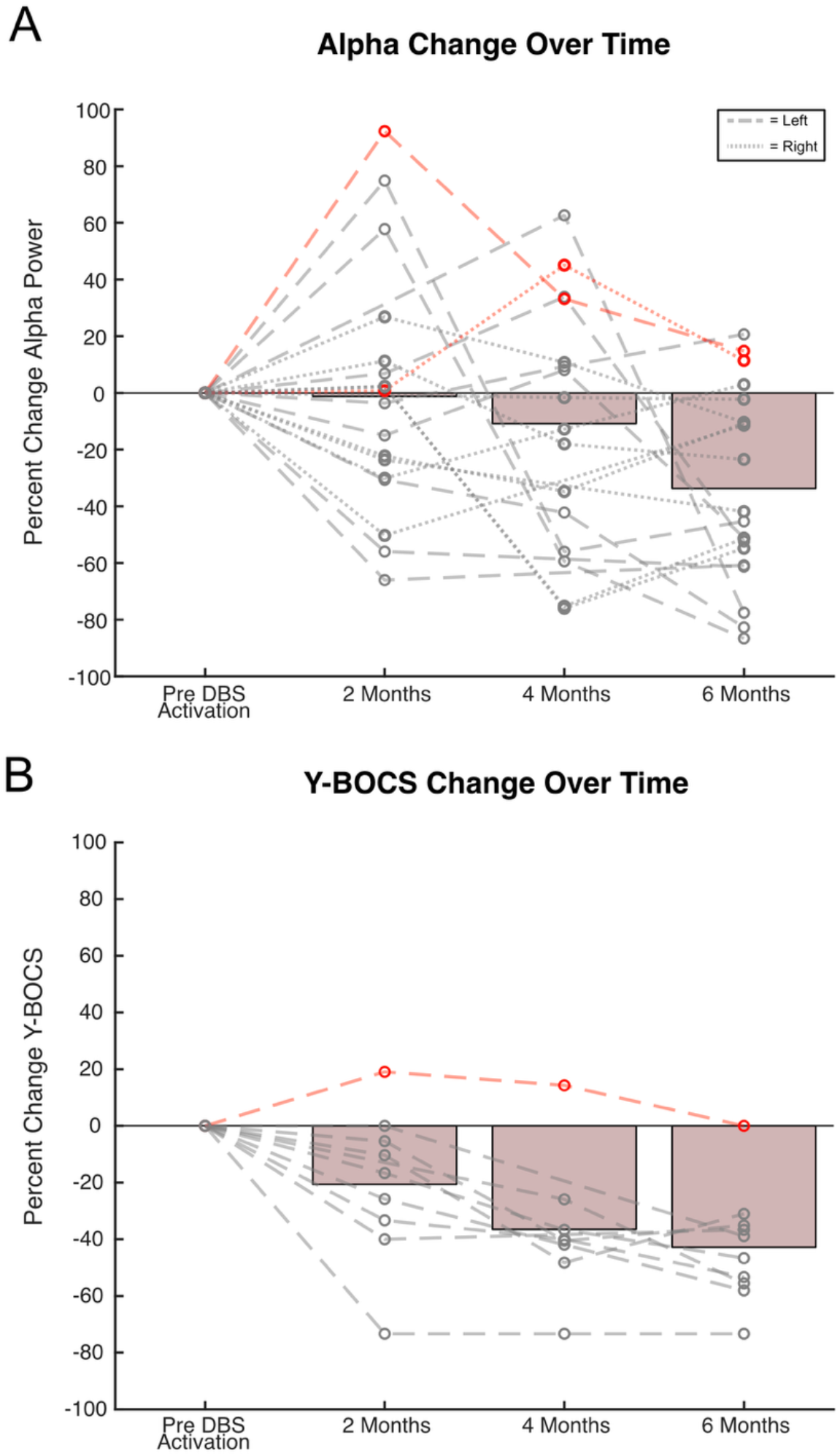
Clinical and Electrophysiological Changes During the Course of Deep Brain Stimulation. **A**) Globus pallidus externus (GPe) alpha power percent change in both left (dashed lines) and right (dotted lines) hemispheres across all available timepoints throughout treatment. **B**) Percent change in the Yale-Brown Obsessive Compulsive Scale (Y-BOCS). The one individual to never exhibit a Y-BOCS decrease (Fig 5B red dotted line) was also the only individual to never exhibit an alpha decrease in either hemisphere (Fig 5A, red dotted line).

## Discussion

ALIC DBS is an effective treatment for severe OCD (Meyer et al., 2024) but there is still variability in treatment outcomes. Developing electrophysiological markers defining a successful treatment effect could help to guide adjustments and improve outcomes (Nho et al., 2024). Our study longitudinally recorded broad frequency spectrum LFP from electrodes anchored in the anterior GPe grey matter, allowing for unique tracking of treatment response in this indirect pathway node. We show that GPe alpha band power decreases significantly during effective therapy, implicating the indirect pathways in DBS treatment mechanisms.

Our findings expand the nascent literature on intracranial markers of OCD and DBS. Comparable to our low frequency alpha changes in DBS for OCD, other studies have implicated low frequency neural activity in OCD DBS (Duffy et al., 2023; Nho et al., 2024; Rappel et al., 2018; Vissani et al., 2023; Xiong et al., 2023). One study combined data from anatomically variable recording sites in the ventral capsule/ventral striatum (VC/VS) and bed nucleus of the stria terminalis (BNST) in 12 OCD patients receiving DBS (Provenza et al., 2024) showing decreased theta/alpha (9+/-2.5 Hz) periodicity in DBS responders. In a case report of VC/VS DBS for OCD, alpha band activity decreased during treatment, with the exception of an increase in alpha activity during a period of worsening symptoms (Vissani et al., 2023). A cross-sectional study of 11 VC/VS DBS patients with recordings across various anatomical sites found an increase in alpha power during provoked mental compulsions, confined to the GPe (Arbab et al., 2025). While prior studies utilized different brain sites and methods of recording and DBS targeting, taken together this prior literature suggests two hypotheses. First, that alpha power may be an oscillatory measure of OCD treatment response. Second, that the GPe may be the site of most clinical relevance for physiological monitoring. Our work unites these two hypotheses by demonstrating alpha changes while recording in the GPe longitudinally.

A strength of our approach is the anatomical specificity achieved by combining prospective tractographic white matter targeting with microelectrode grey matter confirmation, allowing us to consistently capture LFP recordings from the anterior (limbic-cognitive) GPe territory. In addition, collecting resting state data in a controlled laboratory setting allowed us to take full advantage of the Medtronic Percept’s ability to record for extended periods of time. We recorded 5 minutes of eyes-open data per visit, providing raw time domain data which would be unavailable from the Percept’s brief (seconds long) data snapshot home functionality. We were therefore able to implement a rigorous artifact clearing and analysis pipeline before examining each canonical frequency band. Future, larger cohorts will be required to further validate and dissect these electrophysiological patterns. Within our limited sample, there was also only a single patient who did not show any clinical improvement over time, illustrating the effectiveness of our treatment strategy. A larger and more densely sampled cohort of similarly implanted OCD DBS patients would presumably provide additional nonresponders for analyses, including machine learning approaches (Alagapan et al., 2023). Beyond our limited sample size, a universal issue in psychiatric DBS studies, another limitation of this study is that the Percept device is not technologically optimized for recording reliable neural data either in the delta or high gamma range (Medtronic, 2021), as opposed to more invasive recording approaches (Scangos et al., 2021).

Using a consistent individualized tractography method to define the DBS target, we observed OCD improvement in 9/10 patients along with alpha activity decreases in anterior GPe. We observed that the only individual whose OCD symptoms never improved over the course of the study was also the only individual whose GPe alpha power never decreased. In future, longer and more frequent longitudinal sampling will be required to obtain finer grained electrophysiological data to assess if this alpha decrease serves as a stable response biomarker and if fluctuations might guide DBS adjustments.

## Data Availability

All data produced in the present study are available upon reasonable request to the authors

## Author Contributions

M.F. and A.H.S. designed the study. B.H.K. performed the DBS surgery. H.N.S. and K.S.C. performed imaging analysis. S.O. and Z.I. collected the data. Z.I., I.S., A.C.W, and A.H.S. performed electrophysiological analysis. Z.I., B.H.K., I.S., H.S.M., K.S.C., A.C.W, M.F., and A.H.S provided feedback for data analysis and interpretation. All authors have approved the manuscript in its final form.

## Acknowledgements

We thank the patients and their families for agreeing to participate in research.

